# Effectiveness of anterior cruciate ligament injury prevention programmes in sport: a scoping review

**DOI:** 10.1101/2022.08.10.22277937

**Authors:** Michele Margelli, Francesco De Mucci, Francesco Nadaletto, Tommaso Marzi Manfroni, Andrea Beccari

## Abstract

**Background:** Anterior cruciate ligament injury is the most common ligament injury of the knee and one of the most serious and common injuries in sport. This injury causes a number of both short-term and longterm negative consequences that negatively impact both sports performance and quality of life in general. The literature has recognised the need to improve primary prevention with the aim of decreasing the ACL injury rate. For this reason, numerous ACL-specific prevention programmes have been proposed in recent years that include various types of exercises and different methods of administration. The main objective of this scoping will be to report what the literature tells us about these programmes and prevention for ACL injuries.

**Inclusion criteria:** Every study about prevention programmes for LCA injury on athletes of all ages participating in professional or amateur sports will be included. This scoping review will consider studies conducted in any context.

**Methods:** The proposed scoping review will be conducted in accordance with the Joanna Briggs Institute methodology (JBI) for scoping reviews.

The search will be carried out on 6 databases: *MEDLINE, Cochrane Central, Scopus, CINHAL, Embase, and PEDro*.

Selection and data extraction will be conducted by two blind independent researchers and inconsistencies will be resolved by a third reviewer.

The results will be presented in a schematic, tabular and descriptive format that will line up with the objectives and scope of the review.

**Conclusions:** This review provides a comprehensive overview of prevention for ACL injuries in sport, in particular the effectiveness of existing prevention programmes. The information provided will be useful for physiotherapists and athletic trainers during the sports season and as a therapeutic education tool. The results of this research will be published in scientific research databases.

## Background

Anterior cruciate ligament injury is the most common knee’s ligament injury^1^. Furthermore, it is one of the most common and serious injury in sport^2 3^

An ACL injury provoques a number of significant consequences, as an extended absence from sporting activities (200 days on average)^4^, that could lead to a complete withdrawal from sport due to the fear of re-injury ^5^. Moreover, there is a high risk of recurrence^6^ and, in addition, a high percentage (35%) of individuals fail to return to their pre-injury performance levels^7 8^. Additionally, there are medium to long term consequences such as chronic instability, secondary meniscus and cartilage damage and osteroarthritis^9 10 11^

There appears to be an increasing focus in the literature on the need for better primary prevention of ACL injuries ^8 12^. Talking about prevention is necessary because the majority of cruciate ligament injuries occur with non-contact dynamics and that many of the main risk factors are modifiable some of them are a specific biomechanical and neuromuscular deficits, high BMI, muscle fatigue, landing and pivot differences, proprioception and core stability ^9^.

There are many prevention programs in the literature that include different types of interventions: neuromuscular and proprioceptive training, stretching, plyometrics, movement training, core strengthening, balance training, resistance training, and speed training ^13^. Studies that have examined the effectiveness of these programs have found great variability in effect, in terms of reducing the risk of ACL injury (from no significant effect to a 62% risk reduction) ^14^. This variability depends on many variables to consider: age of participants, type of sport, professional or nonprofessional athlete, gender, program compositions in terms of dosage and type of intervention.

A 2018 guideline from the Academy of Orthopaedic Physical Therapy and the American Academy of Sports Physical Therapy explain the effectiveness of prevention in relation to some of these variables (age, gender, type of sport, type of intervention) and identified several effective programs in relation to these ones recommending the use of some of these programs to all athletes.^15^

It remains unclear which interventions are most effective in terms of type of exercise or in terms of mode and timing of administration.^16^

The aim of this scoping is to describe what literature presented us so far in the area of primary prevention of cruciate ligament injuries in sport.

### Review question

Which programmes prevent ACL injury in sport and how effective are they? The main objectives of this study will be:

1. Identify and describe LCA injury prevention programmes existing in the literature
2. Describe the effectiveness of these programmes in terms of reducing the injury rate, in relation to the type of sport and the type of athlete (professional or amateur).
3. Identify knowledge gaps on this topic.

## Methods

### Study design and protocol

The proposed scoping review will be conducted in accordance with the Joanna Briggs Institute methodology (JBI) for scoping reviews.

The Preferred Reporting Items for Systematic reviews and Meta-Analyses extension for Scoping Reviews (PRISMA-ScR) Checklist for reporting will be used, and it is priori registered at Medxriv (https://www.medrxiv.org).

### Search strategy

The search will be carried out on 6 databases: MEDLINE, Cochrane Central, Scopus, CINHAL, Embase, and PEDro. Studies will also be included searching in the bibliography of relevant revisions, and in Google Scholar. Further research of Gray literature will be carried out through open gray.eu, a multidisciplinary European database. Keywords inserted are: ACL, prevention, second injury, sport, adult, athlete. As recommended in all JBI types of reviews and PRISMA-S, a three-step search strategy should be developed:

The first step is represented by an initial limited search of an appropriate online database which is relevant to the topic (MEDLINE). This research is based on identifying an appropriate search string by following the acronym PCC (Population, Concept and Context) proposed by The JBI and using the keywords found by the preliminary background search. The initial search is followed by the analysis of the text words contained in the title, of the retrieved papers’ abstract, and of the index terms, used to describe the articles, on SR accelerator (https://sr-accelerator.com).

The second step concerns a search based on all identified keywords and index terms that should be undertaken across all included databases.

The third step, the reference list of identified reports and articles should be searched for additional sources.

The search strategies were peer-reviewed by an experienced librarian and were further refined through team discussion. No search limitations and filters applied (language and time). Reviewers’ intent to contact authors of primary sources or reviews for further information. Complete search strategy for Medline is included as an appendix 1 to the protocol. Search strategy will be adapted to be used in other databases.

### Inclusion criteria

We will follow the acronym PCC to describe the elements of the inclusion criteria:

#### Population

This scoping review will consider studies with athletes of all ages participating in professional or amateur sports.

#### Concept

This scoping review will consider research studies on prevention programmes for anterior cruciate ligament rupture in athletes.

#### Context

This scoping review will consider studies conducted in any environment.

#### Sources

This scoping review will consider any study designs or publication type for inclusion. No date and geographical limits will be used. We will consider articles in English and Italian; or articles of any language, with at least available English abstract. Studies that do not meet the above-stated Population-Concept-Context (PCC) criteria or which provide insufficient information will be excluded. The rational for choice is part of our initial application.

### Study selection

For the selection process, the first thing to do is to select a sample of 25 title/abstracts that will be analyzed from the entire team using eligibility criteria and definitions/elaboration document. Then, the team will discuss discrepancies and make modifications to the eligibility criteria and definitions/elaboration document. The team will only start screening when 75% (or greater) of agreement between reviewers will be achieved.

The selection phase will begin assessing all the titles and abstracts of the studies retrieved using the search strategy and those from additional sources. These studies will be screened independently by two review authors, to identify those that potentially meet the inclusion criteria. Any disagreement will be solved by discussion with a third author. Subsequently, the full text of these potentially eligible studies will be independently assessed for eligibility by the two reviewers. The reasons for excluding articles will be recorded. We developed a google form (charting table) containing the elements to standardize the selection.

There should be a narrative description of the selection process accompanied by a flowchart of review process (from the PRISMA-ScR statement).

### Data extraction

Extraction module (in appendix B) will be reviewed, before the implementation, by the research team and pre-tested to ensure that the form accurately captures the information. Modifications will be detailed in the full scoping review.

Data extraction will be conducted by two blind independent researchers and inconsistencies will be resolved by a third reviewer.

### Data items

Key information will be described in a charting table with the description of: Author; Publication year; Place of study’s conduct; Setting of study’s conduct; Methodology/ type of study; Aims of the study; Population characteristics from which patients are extracted, including gender and age; Concept: training modalities, type of exercises, sets, frequency and total time per session, type of athlete (professional or amateur), type of sport, outcome measures.

### Critical appraisal of individual sources of evidence

Not provided.

### Data management

As a scoping review, the purpose is to aggregate the findings and to present an overview of the research rather than to evaluate the quality of the individual studies. The results will be presented as a map of data, which are extracted from different documents. These will be included in a schematic, tabular and descriptive format that will line up with the objectives and scope of the review. Descriptive analysis will consist on a distribution of the evidence sources by year or period of publication, countries of origin, area of intervention (clinical, political, educational, etc.) and research methods. The results will be presented as: population (gender, age), type of sport, prevention programme or training mode, decrease LCA injury rate. An overall classification of prevention programmes with narrative description and table will be proposed.

## Data Availability

All data produced are available online as indicated in the bibliography

https://pubmed.ncbi.nlm.nih.gov/

https://pedro.org.au/

https://www.ebsco.com/it-it/prodotti/banche-dati-per-la-ricerca/cinahl-database

https://www.embase.com/

https://www.scopus.com/

https://www.cochranelibrary.com/

## Data Availability

Due to the nature of this research, participants of this study did not agree for their data to be shared publicly, so supporting data is not available.

## Appendix A Complete search strategy

(“Anterior Cruciate Ligament”[MeSH Terms] OR “Anterior Cruciate Ligament”[All Fields] OR (“Anterior Cruciate Ligament”[MeSH Terms] OR (“anterior”[All Fields] AND “cruciate”[All Fields] AND “ligament”[All Fields]) OR “Anterior Cruciate Ligament”[All Fields] OR “acl”[All Fields])) AND (“prevention and control”[MeSH Subheading] OR “prevention and control”[All Fields] OR “preventi*”[All Fields] OR “recurren*”[All Fields] OR (“recidivate”[All Fields] OR “recidivated”[All Fields] OR “recidivating”[All Fields] OR “Recidivism”[MeSH Terms] OR “Recidivism”[All Fields] OR “recidivisms”[All Fields]) OR “Recidivism”[MeSH Terms] OR (“reinjuries”[MeSH Terms] OR “reinjuries”[All Fields] OR “reinjury”[All Fields]) OR “Recurrence”[MeSH Terms] OR (“recurrance”[All Fields] OR “Recurrence”[MeSH Terms] OR “Recurrence”[All Fields] OR “recurrences”[All Fields] OR “recurrencies”[All Fields] OR “recurrency”[All Fields] OR “recurrent”[All Fields] OR “recurrently”[All Fields] OR “recurrents”[All Fields]) OR “second injury”[All Fields] OR “second tears”[All Fields] OR “Secondary Prevention”[MeSH Terms] OR “Secondary Prevention”[All Fields] OR (“Secondary Prevention”[MeSH Terms] OR (“secondary”[All Fields] AND “prevention”[All Fields]) OR “Secondary Prevention”[All Fields] OR (“early”[All Fields] AND “therapy”[All Fields]) OR “early therapy”[All Fields]) OR ((“injurie”[All Fields] OR “injuried”[All Fields] OR “injuries”[MeSH Subheading] OR “injuries”[All Fields] OR “wounds and injuries”[MeSH Terms] OR (“wounds”[All Fields] AND “injuries”[All Fields]) OR “wounds and injuries”[All Fields] OR “injurious”[All Fields] OR “injury s”[All Fields] OR “injuryed”[All Fields] OR “injurys”[All Fields] OR “injury”[All Fields]) AND (“risk reduction behavior”[MeSH Terms] OR (“risk”[All Fields] AND “reduction”[All Fields] AND “behavior”[All Fields]) OR “risk reduction behavior”[All Fields] OR (“risk”[All Fields] AND “reduction”[All Fields]) OR “risk reduction”[All Fields] OR “adaptation, psychological”[MeSH Terms] OR (“adaptation”[All Fields] AND “psychological”[All Fields]) OR “psychological adaptation”[All Fields] OR (“risk”[All Fields] AND “reduction”[All Fields]))) OR (“prevent”[All Fields] OR “preventability”[All Fields] OR “preventable”[All Fields] OR “preventative”[All Fields] OR “preventatively”[All Fields] OR “preventatives”[All Fields] OR “prevented”[All Fields] OR “preventing”[All Fields] OR “prevention and control”[MeSH Subheading] OR (“prevention”[All Fields] AND “control”[All Fields]) OR “prevention and control”[All Fields] OR “prevention”[All Fields] OR “prevention s”[All Fields] OR “preventions”[All Fields] OR “preventive”[All Fields] OR “preventively”[All Fields] OR “preventives”[All Fields] OR “prevents”[All Fields]) OR “relapse preventions”[All Fields] OR ((“Anterior Cruciate Ligament”[MeSH Terms] OR (“anterior”[All Fields] AND “cruciate”[All Fields] AND “ligament”[All Fields]) OR “Anterior Cruciate Ligament”[All Fields] OR “acl”[All Fields]) AND (“reinjuries”[MeSH Terms] OR “reinjuries”[All Fields] OR (“second”[All Fields] AND “injury”[All Fields]) OR “second injury”[All Fields]))) AND (“Sports”[MeSH Terms] OR (“sport s”[All Fields] OR “Sports”[MeSH Terms] OR “Sports”[All Fields] OR “sport”[All Fields] OR “sporting”[All Fields]) OR “athlet*”[All Fields] OR “Athletes”[MeSH Terms] OR “Soccer”[MeSH Terms] OR (“Soccer”[MeSH Terms] OR “Soccer”[All Fields] OR “soccers”[All Fields]) OR “Football”[MeSH Terms] OR (“Football”[MeSH Terms] OR “Football”[All Fields] OR “football s”[All Fields] OR “footballer”[All Fields] OR “footballer s”[All Fields] OR “footballers”[All Fields] OR “footballs”[All Fields]) OR “Basketball”[MeSH Terms] OR (“basket”[All Fields] OR “basketing”[All Fields] OR “baskets”[All Fields]))

## REFERENCE

1. Holly J Silvers-Granelli, Mario Bizzini, Amelia Arundale, Bert R Mandelbaum, Lynn Snyder-Mackler Does the FIFA 11+ Injury Prevention Program Reduce the Incidence of ACL Injury in Male Soccer Players? Clin Orthop Relat Res. 2017 Oct;475(10):2447–2455. doi: 10.1007/s11999-017-5342-5.

2. Amelia J H Arundale, Holly J Silvers-Granelli, Grethe Myklebust ACL injury prevention: Where have we come from and where are we going? J Orthop Res. 2022 Jan;40(1):43–54. doi: 10.1002/jor.25058. Epub 2021 May 11.

3. Alicia M Montalvo, Daniel K Schneider, Paula L Silva, Laura Yut, Kate E Webster, Michael A Riley, Adam W Kiefer, Jennifer L Doherty-Restrepo, Gregory D Myer ‘What’s my risk of sustaining an ACL injury while playing football (soccer)?’ A systematic review with meta-analysis. Br J Sports Med. 2019 Nov;53(21):1333–1340. doi: 10.1136/bjsports-2016-097261. Epub 2018 Mar 29.

4. Raouf Nader Rekik, Montassar Tabben, Cristiano Eirale, Philippe Landreau, Rachid Bouras, Mathew G Wilson, Scott Gillogly, Roald Bahr, Karim Chamari ACL injury incidence, severity and patterns in professional male soccer players in a Middle Eastern league. BMJ Open Sport & Exercise Medicine 2018;4:e000461. doi:10.1136/bmjsem-2018-000461

5. Stephanie R Filbay, Hege Grindem Evidence-based recommendations for the management of anterior cruciate ligament (ACL) rupture. Best Pract Res Clin Rheumatol. 2019 Feb;33(1):33–47. doi: 10.1016/j.berh.2019.01.018. Epub 2019 Feb 21.

6. George J Davies, Eric McCarty, Matthew Provencher, Robert C Manske ACL Return to Sport Guidelines and Criteria. Curr Rev Musculoskelet Med. 2017 Sep;10(3):307–314. doi: 10.1007/s12178-017-9420-9.

7. Nicky van Melick, Robert E H van Cingel, Frans Brooijmans, Camille Neeter, Tony van Tienen, Wim Hullegie, Maria W G Nijhuis-van der Sanden Evidence-based clinical practice update: practice guidelines for anterior cruciate ligament rehabilitation based on a systematic review and multidisciplinary consensus. Br J Sports Med. 2016 Dec;50(24):1506–1515. doi: 10.1136/bjsports-2015-095898. Epub 2016 Aug 18

8. Markus Waldén, Martin Hägglund, Henrik Magnusson, Jan Ekstrand ACL injuries in men’s professional football: a 15-year prospective study on time trends and return-to-play rates reveals only 65% of players still play at the top level 3 years after ACL rupture. Br J Sports Med 2016;50:744–750. doi:10.1136/bjsports-2015-095952

9. Rafael J Acevedo, Alexandra Rivera-Vega, Gerardo Miranda, William Micheo Anterior cruciate ligament injury: identification of risk factors and prevention strategies. Curr Sports Med Rep. May-Jun 2014;13(3):186–91. doi: 10.1249/JSR.0000000000000053.

10. Bing Yu, William E Garrett Mechanisms of non-contact ACL injuries. Br J Sports Med. 2007 Aug;41 Suppl 1(Suppl 1):i47–51. doi: 10.1136/bjsm.2007.037192.

11. Timothy E Hewett, Gregory D Myer, Kevin R Ford, Mark V Paterno, Carmen E Quatman Mechanisms, prediction, and prevention of ACL injuries: Cut risk with three sharpened and validated tools. J Orthop Res. 2016 Nov;34(11):1843–1855. doi: 10.1002/jor.23414. Epub 2016 Sep 19.

12. Barry P. Boden, Frances T. Sheehan, Joseph S. Torg, and Timothy E. Hewett Non-contact ACL Injuries: Mechanisms and Risk Factors. J Am Acad Orthop Surg. 2010 September ; 18(9): 520–527.

13. Joel J. Gagnier, Hal Morgenstern, Laura Chess Interventions designed to prevent anterior cruciate ligament injuries in adolescents and adults: a systematic review and meta-analysis. Am J. Sports Med. 2013 Aug; 41(8):1952–62. doi: 10.1177/0363546512458227. Epub 2012 Sep 12.

14. Yu-Lun Huang, Jaehun Jung, Colin M S Mulligan, Jaekeun Oh, Marc F Norcross A Majority of Anterior Cruciate Ligament Injuries Can Be Prevented by Injury Prevention Programs: A Systematic Review of Randomized Controlled Trials and Cluster-Randomized Controlled Trials With Meta-analysis. Am J Sports Med. 2020 May; 48(6):1505–1515. doi: 10.1177/0363546519870175. Epub 2019 Aug 30.

15. Amelia J H Arundale, Mario Bizzini, Airelle Giordano, Timothy E Hewett, David S Logerstedt, Bert Mandelbaum, David A Scalzitti, Holly Silvers-Granelli, Lynn Snyder-Mackler Exercise-Based Knee and Anterior Cruciate Ligament Injury Prevention. J Orthop Sports Phys Ther.2018 Sep; 48(9):A1–A42. doi: 10.2519/jospt.2018.0303.

16. Jeffrey B Taylor, Justin P Waxman, Scott J Richter, Sandra J Shultz Evaluation of the effectiveness of anterior cruciate ligament injury prevention programme training components: a systematic review and meta-analysis. Br J Sports Med 2015;49:79–87.

